# Efficacy of Preemptive Intravenous Ibuprofen and Dexketoprofen on Postoperative Opioid Consumption in Laparoscopic Cholecystectomy: Randomized Controlled Study

**DOI:** 10.1101/2025.01.12.25320436

**Authors:** Celaleddin Soyalp, Ahmet Murat Yayik, Ersoy Öksüz, Nurettin Yüzkat

**Affiliations:** Department of Anaesthesiology and Reanimation, Van Yuzuncu Yil University School of Medicine, Van, Turkey; Department of Anaesthesiology and Reanimation, Atatürk University, School of Medicine, Erzurum, Turkey; Department of medical pharmacology, malatya turgut ozal university, School of Medicine, Malatya, Turkey

**Author notes:** Corresponding Author: Ahmet Murat Yayik, MD, Address 1: Ataturk University School of Medicine, Department of Anesthesiology and Reanimation, 25070, Erzurum/ Turkey, Address 2: Clinical Research, Development and Design Application and Research Center, Ataturk University School of Medicine, 25240, Erzurum, Turkey. **Funding information** The authors did not receive any funds, grants, or other supports. **Declaration of competing interest** All authors have completed the ICMJE uniform disclosure form. The authors have no con flicts of interest to declare. **Use of AI for Writing Assistance:** None.

**Keywords:** Laparoscopic cholecystectomy, postoperative preemptive analgesia, dexketoprofen, ibuprofen

## Abstract

**Background:** To compare the effects of preemptive single-dose intravenous (IV) ibuprofen and dexketoprofen on postoperative pain and opioid consumption in patients undergoing laparoscopic cholecystectomy (LCC).

**Methods:** The study included 90 patients aged 18-65 years with an ASA score of I or II who underwent LCC. Patients were equally divided into three groups: Control Group (Group P), 100 cc 0.9% NaCl was infused intravenously over 30 min, Dexketoprofen Group (Group D), 50 mg dexketoprofen in 100 cc 0.9% NaCl was infused intravenously over 30 min, and Ibuprofen Group (Group I), 800 mg ibuprofen in 100 cc 0.9% NaCl was administered intravenously over 30 min. Visual Analog Scale (VAS) scores and opioid requirement were recorded at 1, 2, 4, 6, 12 and 24 hours postoperatively.

**Results:** There was no significant difference between the Dexketoprofen and Ibuprofen groups with regard to VAS scores, whereas VAS scores were higher in the control group than in other groups. In addition, fentanyl consumption was higher in the control group at 0-6 hours and at 24 hours compared to the other two groups.

**Conclusion:** Preemptive ibuprofen and dexketoprofen administration showed no significant difference with regard to postoperative pain and opioid consumption, particularly within the first 6 hours after LCC.

## INTRODUCTION

Laparoscopic cholecystectomy (LCC) is the most commonly performed surgical method in the treatment of gallstones. LCC is highly popular among gallstone patients mainly because it provides early mobilization as well as reduced pain compared to open surgery. However, postoperative pain has been reported in many patients after LCC. ^[1]^ The intensity of pain is usually mild to moderate. ^[2]^ Nevertheless, in some cases, severe pain develops, which may lead to prolonged recovery time and chronic pain.^[3]^ Pain occurring after LCC is often caused by tissue trauma, abdominal distension, chemical irritation of the peritoneum, and irritation of the diaphragm due to carbon dioxide (CO_2_) dissolution in the abdomen. ^[4]^

Opioid analgesics and non-steroidal anti-inflammatory drugs (NSAIDs) are widely used preoperatively to prevent or reduce postoperative pain in both open surgery and laparoscopic surgery. In addition to their analgesic effects, these drugs also have critical roles in the recovery of patients. ^[5]^ Nevertheless, opioid analgesics have limited clinical use since they do not have any role in the improvement of the pathophysiological condition induced by the pain and they are known to cause various side effects. For this reason, combined use of opioid drugs and NSAIDs has recently become a popular technique for postoperative analgesia. ^[4,6,7]^ This technique reduces the use and side effects of high-dose opioid administration and also provides effective analgesia. ^[4, 8, 9]^

Dexketoprofen is a non-selective NSAID of the aryl-propionic acid group and is widely used for preemptive analgesia. ^[8]^ Dexketoprofen, like other NSAIDs, produces its activity by inhibiting cyclooxygenase enzymes (COX-1, COX-2). COX is the enzyme that catalyzes the first rate-limiting step in the synthesis of prostaglandins that cause pain and fever and convert arachidonic acid into prostaglandin G2. ^[4,9, 10]^ Dexketoprofen is widely used in daily life as well as in postoperative analgesia and provides short-term analgesic efficacy. ^[8]^

Ibuprofen, in a similar way to dexketoprofen, is an NSAID derived from propionic acid. Its oral form has been widely used in daily life for long years as due to its analgesic, antipyretic, and anti-inflammatory activity. ^[4, 8, 9]^ In 2009, the use of intravenous (IV) ibuprofen as an analgesic was approved by the American Food and Drug Administration (FDA). ^[10, 11]^ Since then, IV ibuprofen has been widely used alone or in combination with opioid drugs for postoperative analgesia. ^[12, 13]^. In previous studies, ibuprofen has been shown to provide safe and effective postoperative analgesia for mild to moderate pain in patients undergoing surgeries such as orthopedic, abdominal, gynecological surgery and LCC. ^[2, 4]^. Additionally, studies comparing the effects of IV ibuprofen and other analgesic drugs on postoperative pain levels have reported that IV ibuprofen provides more effective analgesia compared to other drugs. ^[4]^

The present study aimed to compare the effects of single-dose administration of IV ibuprofen and dexketoprofen on postoperative pain and opioid consumption in patients undergoing LCC.

## MATERIALS AND METHODS

### Ethics Approval

The local institutional review board of the Ethical Committee for Clinical Research of the Medical Faculty of Yüzüncü Yıl University approved the trial (Date: 28.03.2018, Approval No: 04). It conformed to the tenets of the Declaration of Helsinki and was registered before patient enrolment. The study was registered on ClinicalTrials.gov (Identifier: NCT03607266). This randomized-controlled study was designed and conducted according to the guidelines of the Consolidated Standards of Reporting Trials (CONSORT). Written informed consent was obtained from all patients.

### Study population

The study included 90 patients aged 18-65 years with an ASA score of I or II who underwent LCC in Yüzüncü Yıl University Medical School General Surgery Clinic between August 2018 and May 2020. Exclusion criteria were as follows: conversion to open cholecystectomy, ASA score III and IV, chronic diseases such as kidney disease, cancer, heart disease, diabetes, and being followed up in intensive care unit (ICU) after the surgery. All patients underwent physical examination and laboratory investigations the day before the surgery. Patients were asked to indicate the severity of their pain on Visual Analog Scale (VAS), in which 0 indicated no pain and 10 showed the most severe pain.

### Study protocol

The 90 patients included in the study were randomized and equally divided into three groups Control Group (Group P), 100 cc 0.9% NaCl was infused intravenously over 30 min, Dexketoprofen Group (Group D), 50 mg dexketoprofen in 100 cc 0.9% NaCl was infused intravenously over 30 min, and Ibuprofen Group (Group I), 800 mg ibuprofen in 100 cc 0.9% NaCl was administered intravenously over 30 min. Surgical durations and hemodynamic parameters including systolic blood pressure (SBP), diastolic blood pressure (DBP), mean arterial pressure (MAP), oxygen saturation (SpO2), and heart rate (HR) were recorded at certain time intervals.

General anesthesia induction was performed in the same way in all three groups, using IV 2 mg/kg propofol, 2 mcg/kg fentanyl, and 0.6 mg/kg rocuronium. Anesthesia was maintained with 8% desflurane, 40% O_2_, and 60% dry air. When MAP or pulse rate increased by 20% compared to baseline values, 1 mcg/kg fentanyl was administered. Throughout the LCC procedure, intra-abdominal pressure was maintained at 12-14 mmHg in all groups. In each patient, a total of four trocars were inserted in the abdomen, with one of them inserted through the umbilicus, one below the xiphoid process through a 1 cm incision, one through the right anterior axillary line incision, and the remaining one through a 0.5-cm incision on the midclavicular line. After the surgery, local anesthetic infiltration with 5 ml of 0.5% bupivacaine was applied to each trocar entry site. All the surgeries were performed by the same surgical team using the same technique.

At the end of the surgery, IV 0.015 mg/kg atropine and 0.04 mg/kg neostigmine were administered to achieve decurarization. Patients were transferred to the recovery room and then those with a Modified Aldrete Score of 9 and above were transferred to the general ward.

### Postoperative Analgesia

Intravenous patient-controlled analgesia (IV PCA) was set to a bolus dose 25 μg fentanyl without basal infusion, lockout period 10 min, and 6 doses per hour. VAS scores of the patients were recorded while the patients were at rest at 1, 2, 4, 6, 12 and 24 hours postoperatively by the nurse/physician who was unaware of the drugs used for analgesia and the patient groups. Patients with a VAS score ≥ 4 were administered IV 50 mg tramadol in 100 cc saline and the times of administration were recorded. All patients were monitored for side effects such as nausea-vomiting, dry mouth, itching, palpitations, and headache that occurred within the first 24 hours after surgery. At the end of the study, the patients were asked to evaluate their postoperative pain or other discomfort on a three-point scale (1 = bad, 2 = average, 3 = good) and the results were recorded for each patient.

### Power Analysis

The primary aim of the study was to compare fentanyl consumption among the three groups 24 hours postoperatively. To determine the required sample size, a preliminary study was performed with 30 patients. While the mean fentanyl consumption was 192.5 ± 72.7 µg in Group I (n = 10), it was 310.00 ± 123.71 µg in Group D (n = 10), and 317.5 ± 136.95 µg in Group P (n = 10). For total opioid consumption, a sample size of 66 was calculated using GPower (version 3.1.9.2, Dusseldorf, Germany) with an alpha probability of 0.05, a power of 0.95, and a medium-large effect size (0.50). Considering possible dropouts, we included 30 patients in each group to attain higher power for a total of 90 patients.

### Statistical analysis

Data were analyzed using SPSS 25.0 (Armonk, NY: IBM Corp.) and PAST 3 (Hammer, Ø., Harper, D.A.T., Ryan, P.D. 2001. Paleontological statistics). Normal distribution of univariate data was evaluated using the Shapiro-Wilk Francia test and the homogeneity of variance was evaluated using Levene’s test. Normal distribution of multivariate data was evaluated using Mardia’s coefficient and Dornik and Hansen omnibus test and the homogeneity of variance was assessed using the Box’s M test. Two or more groups were compared using One-Way ANOVA (Robust Test: Brown-Forsythe) for parametric data and Kruskal-Wallis H Test with Monte Carlo Simulation for non-parametric data, followed by post-hoc Dunn’s Test. Categorical variables were compared using Pearson’s Chi-Square and Fisher-Freeman-Holton test with Monte Carlo Simulation. Continuous variables were expressed as mean ± standard deviation (SD) and median (minimum / maximum) and categorical variables were expressed as frequencies (n) and percentages (%). A *p* value of <0.05 was considered significant.

## RESULTS

A hundred-three patients were evaluated for enrollment in the study. After excluding 13 patients, others were randomly placed in three groups with 30 patients. Figure 1 represents the CONSORT flow diagram of participants.

**Figure. 1.**
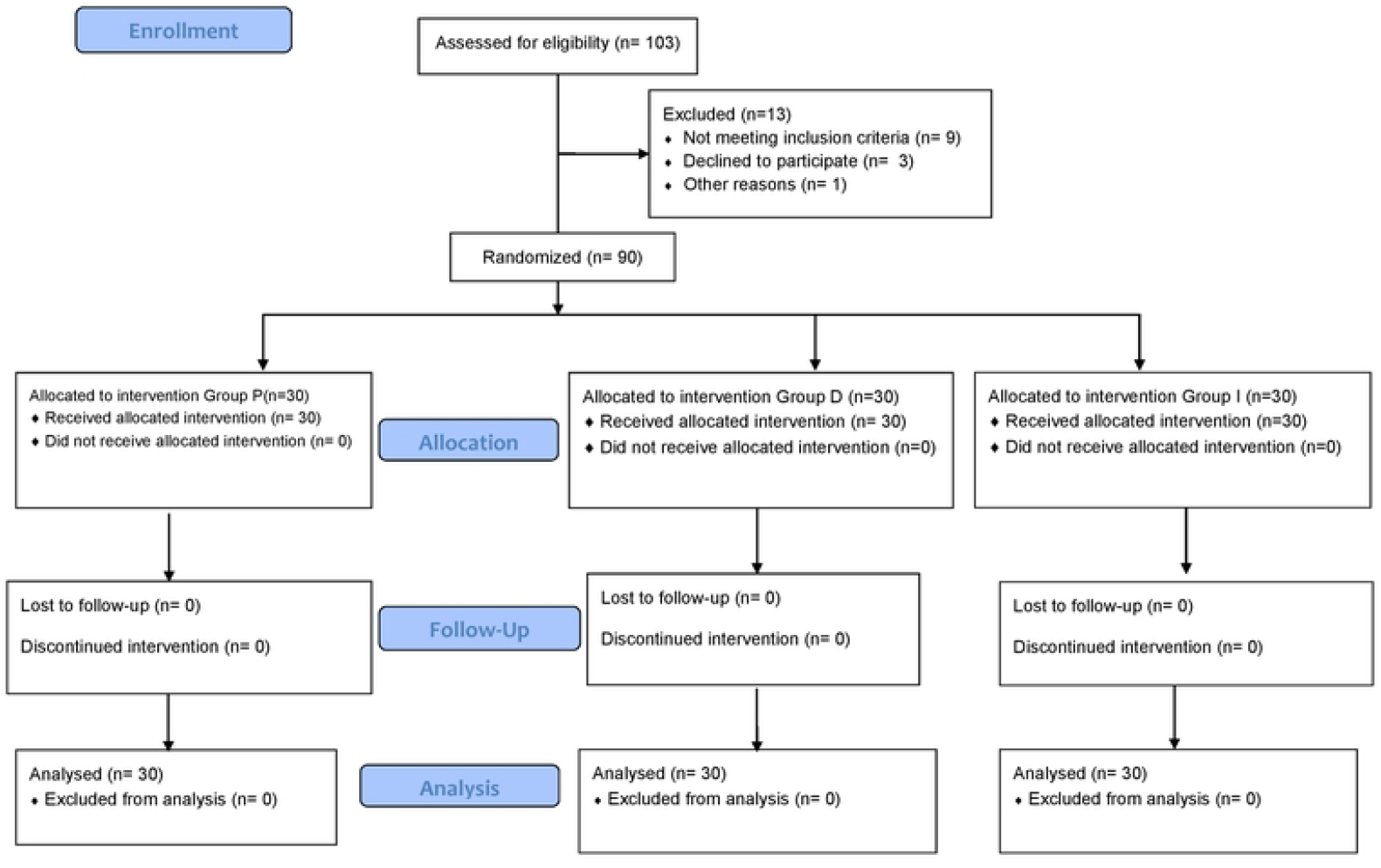

Table 1 presents the demographic characteristics of the patients. No significant difference was observed among the three groups with regard to age, gender, body weight, body height, ASA score, and surgical duration (*p*>0.05 for all).

**Table 1.** Demographic characteristics.

|  | Group I | Group D | Group P | P |
| --- | --- | --- | --- | --- |
|  | (n=30) | (n=30) | (n=30) |  |
| Gender (F/M) | 17/13 | 18/12 | 15/15 | 0.810 <sup>pm</sup> |
| ASA I/II | 12/18 | 11/19 | 14/16 | 0.806 <sup>pm</sup> |
|  | Mean±SD | Mean±SD | Mean±SD |  |
| Age (years) | 45.47±11.32 | 48.10±11.84 | 45.7±12.07 | 0.632 <sup>o</sup> |
| Weight (kg) | 77.93±10.22 | 79.63±14.21 | 81.57±11.07 | 0.503 <sup>o</sup> |
| Height (cm) | 166.37±9.91 | 168.27±10.52 | 168.47±8.86 | 0.658 <sup>o</sup> |
| BMI | 28.20±3.20 | 28.06±3.95 | 28.90±4.45 | 0.671 <sup>o</sup> |
|  | Median (Min/Max) | Median (Min/Max) | Median (Min/Max) |  |
| Surgical duration (min) | 40 (25 / 80) | 45 (20 / 80) | 40 (25 / 75) | 0.132 <sup>kw</sup> |

SD: Standard Deviation, Min.: Minimum, Max.: Maximum. Data is presented as mean±SD <sup>o</sup>
One-Way ANOVA (Robuts Statistic: Brown-Forsythe), <sup>kw</sup> Kruskal-Wallis Test (Monte Carlo), <sup>pm</sup> Pearson Chi-Square Test (Monte Carlo),

There was no significant difference between Group I and D with regard to VAS scores at all times (*p*>0.05). VAS scores were significantly higher in Group P than in other two groups at all times (*p*<0.05) (Table 2).

**Table 2.**
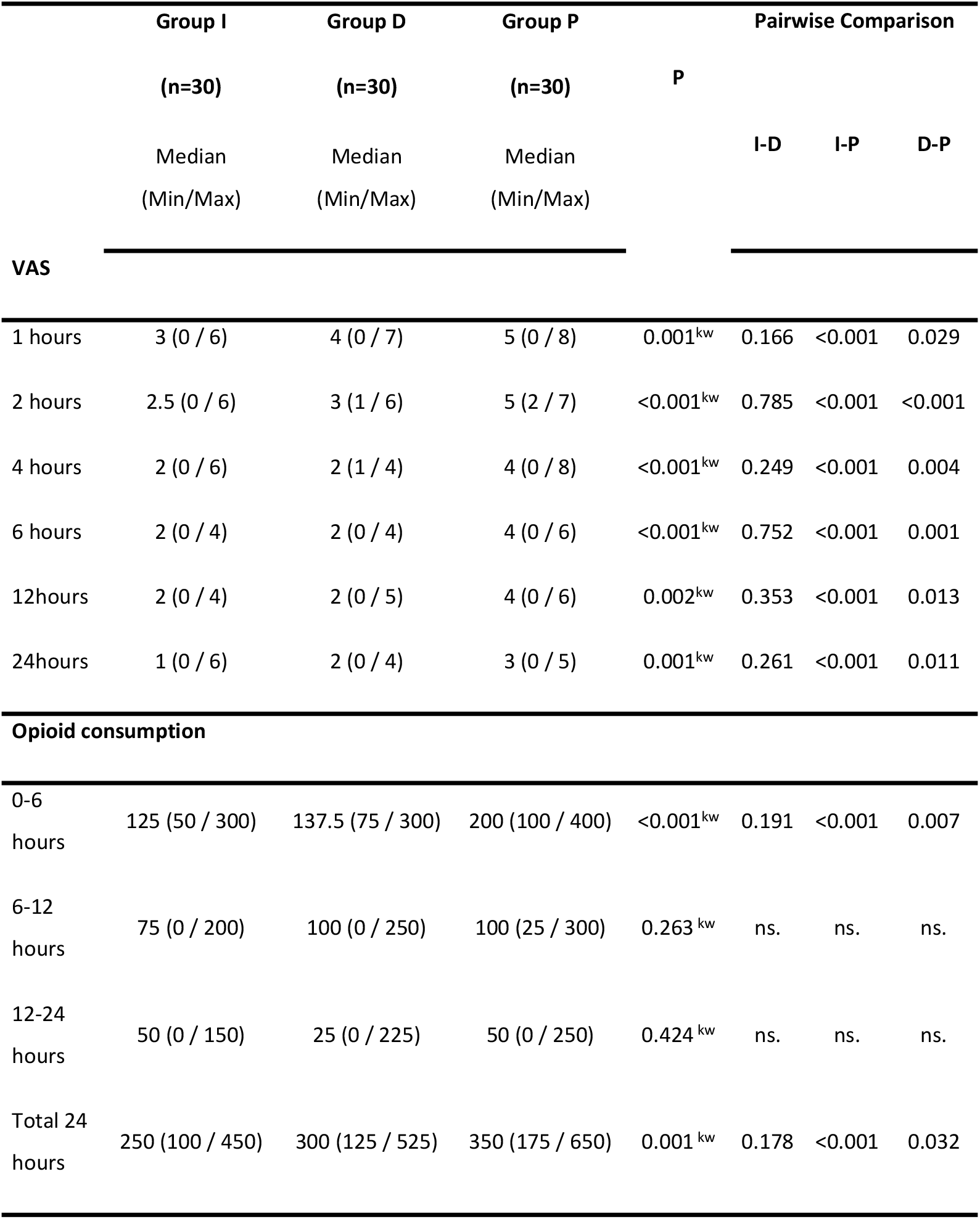
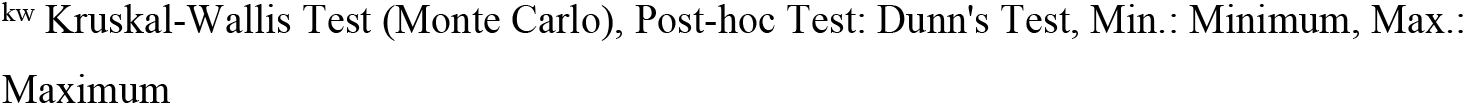
Postoperative Visual Analog Scale Scores and Fentanyl Consumptions.

|  | **Group I** | **Group D** | **Group P** |  | **Pairwise Comparison** |  |  |
| --- | --- | --- | --- | --- | --- | --- | --- |
|  | **(n=30)** | **(n=30)** | **(n=30)** | **P** |  |  |  |
|  | Median | Median | Median |  | **I-D** | **I-P** | **D-P** |
|  | (Min/Max) | (Min/Max) | (Min/Max) |  |  |  |  |
| **VAS** |  |  |  |  |  |  |  |
| 1 hours | 3 (0 / 6) | 4 (0 / 7) | 5 (0 / 8) | 0.001<sup>kw</sup> | 0.166 | <0.001 | 0.029 |
| 2 hours | 2.5 (0 / 6) | 3 (1 / 6) | 5 (2 / 7) | <0.001<sup>kw</sup> | 0.785 | <0.001 | <0.001 |
| 4 hours | 2 (0 / 6) | 2 (1 / 4) | 4 (0 / 8) | <0.001<sup>kw</sup> | 0.249 | <0.001 | 0.004 |
| 6 hours | 2 (0 / 4) | 2 (0 / 4) | 4 (0 / 6) | <0.001<sup>kw</sup> | 0.752 | <0.001 | 0.001 |
| 12hours | 2 (0 / 4) | 2 (0 / 5) | 4 (0 / 6) | 0.002<sup>kw</sup> | 0.353 | <0.001 | 0.013 |
| 24hours | 1 (0 / 6) | 2 (0 / 4) | 3 (0 / 5) | 0.001<sup>kw</sup> | 0.261 | <0.001 | 0.011 |
| **Opioid consumption** |  |  |  |  |  |  |  |
| 0-6 hours | 125 (50 / 300) | 137.5 (75 / 300) | 200 (100 / 400) | <0.001<sup>kw</sup> | 0.191 | <0.001 | 0.007 |
| 6-12 hours | 75 (0 / 200) | 100 (0 / 250) | 100 (25 / 300) | 0.263<sup>kw</sup> | ns. | ns. | ns. |
| 12-24 hours | 50 (0 / 150) | 25 (0 / 225) | 50 (0 / 250) | 0.424<sup>kw</sup> | ns. | ns. | ns. |
| Total 24 hours | 250 (100 / 450) | 300 (125 / 525) | 350 (175 / 650) | 0.001<sup>kw</sup> | 0.178 | <0.001 | 0.032 |

<sup>kw</sup> Kruskal-Wallis Test (Monte Carlo), Post-hoc Test: Dunn's Test, Min.: Minimum, Max.: Maximum

Postoperative fentanyl consumption was assessed at 0-6 hours, 6-12 hours, 12-24 hours, and 24 hours. Fentanyl consumption was significantly higher in Group P than in Group D and I both at 0-6 hours (200, 137.5, and 125 mcg, respectively, *p*<0.05) and at 24 hours (350, 300, and 250 mcg, respectively, *p*<0.05). However, there was no significant difference among the three groups with regard to fentanyl consumption at 6-12 and 12-24 hours (*p*>0.05). Similarly, there was no significant difference between Group I and D in terms of fentanyl consumption at all times (*p*>0.05) (Table 2).

Intraoperative hemodynamic parameters were compared at three time points: before the incision, five min after the incision, and after the completion of the surgical procedure. There was no significant difference among the three groups with regard to all three time points (*p*>0.05) (Table 3).

**Table 3.** Hemodynamic Parameters.

|  | **Group I** | **Group D** | **Group P** | **P** |
| --- | --- | --- | --- | --- |
|  | **(n=30)** | **(n=30)** | **(n=30)** |  |
|  | Median | Median | Median |  |
|  | (Min/Max) | (Min/Max) | (Min/Max) |  |
| **Heart rate** |  |  |  |  |
| Pre-incision | 77.5 (59 / 98) | 77 (61 / 110) | 81.5 (60 / 93) | 0.118<sup>kw</sup> |
| 5 min. after incision | 76.75 (50 / 90) | 73.5 (61 / 108) | 81 (64.5 / 98) | 0.177<sup>kw</sup> |
| End of surgery | 86 (47 / 97) | 77 (54 / 140) | 80 (56 / 100) | 0.192<sup>kw</sup> |
| **Systolic blood pressure** |  |  |  |  |
| Pre-incision | 140 (100 / 177) | 131 (90 / 180) | 140 (110 / 180) | 0.427<sup>kw</sup> |
| 5 min. after incision | 129.5 (89 / 153) | 131 (90 / 151) | 129.5 (96 / 159) | 0.986<sup>kw</sup> |
| End of surgery | 132.5 (114 / 175) | 130.5 (90 / 152) | 130.5 (84 / 161) | 0.564<sup>kw</sup> |
| **Diastolic blood pressure** |  |  |  |  |
| Pre-incision | 78 (57 / 104) | 75.5 (60 / 102) | 76.5 (61 / 104) | 0.694<sup>kw</sup> |
| 5 min. after incision | 79 (56 / 91) | 75.25 (58 / 90.5) | 77.5 (62 / 90.5) | 0.404<sup>kw</sup> |
| End of surgery | 80.5 (59 / 109) | 80 (52 / 104) | 84.5 (68 / 99) | 0.187<sup>kw</sup> |

<sup>kw</sup> Kruskal-Wallis Test (Monte Carlo), Post Hoc Test: Dunn's Test, Min: Minimum, Max.: Maximum

Postoperative rescue analgesia was administered to 13 patients in Group P, 8 patients in Group D, and 7 patients in Group I and no significant difference was found among the three groups (*p*=0.202). Similarly, there was no significant difference among the three groups with regard to the incidence of opioid-induced nausea, vomiting, and dry mouth (*p*>0.05) and in terms of postoperative patient satisfaction (*p*=0.534) (Table 4).

**Table 4.**
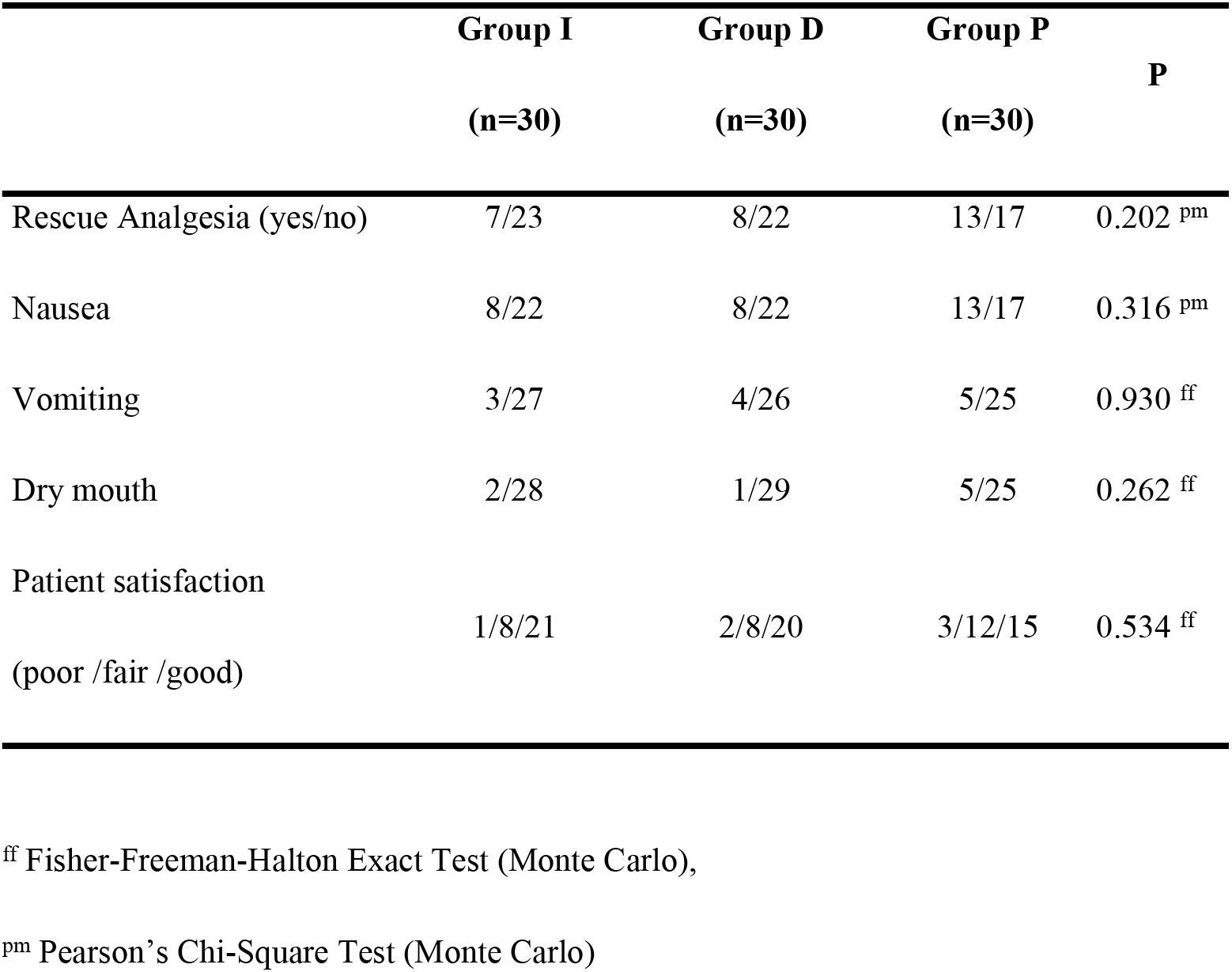
Opioid-induced side effects and patient satisfaction.

## DISCUSSION

The findings indicated that that preemptive single-dose IV ibuprofen and dexketoprofen administration reduced the pain scores and opioid consumption after LCC, particularly in the early postoperative period. However, ibuprofen and dexketoprofen were found to have no superiority over each other.

Studies have shown that postoperative pain is commonly seen after LCC and that the VAS scores of the patients range between 3 and 5. ^[2, 4]^ Pain after LCC is often seen in the first hours of the postoperative period and the intensity of pain usually peaks during this period. The pain then decreases considerably in the following hours and usually disappears within 2-3 days ^2^. In our study, the VAS scores of the patients in the control group were similar to those reported in the literature. ^[14]^ In this group, mean VAS score was between 3 and 5 at 24 hours, while it was 5 in the first 2 hours and gradually decreased to 3 after 6 hours.

Studies conducted on patients undergoing LCC surgery reported that the postoperative VAS scores and opioid consumption rates of patients that received a preemptive 400 mg single dose of IV ibuprofen were lower than those of control group. Moreover, the frequencies of additional analgesia and postoperative rescue analgesia were also found to be lower in these patients than in control subjects. ^[1]^ In a double-blind placebo-controlled randomized study conducted in abdominal surgery patients, it was found that both the pain intensity and morphine use in the postoperative period were significantly lower in patients that received 800 mg IV ibuprofen in the preoperative period compared to the placebo group ^4^. In another study, the use of a single dose of IV 800 mg ibuprofen in various surgeries such as septorhinoplasty and thyroid surgery was similar to those of other studies. ^[15]^

Among the studies comparing the effects of preemptive use of ibuprofen and other NSAIDs on postoperative pain, numerous studies reported that IV ibuprofen was more effective than other NSAIDs. In one of these studies, the authors evaluated patients that underwent laparoscopic gastrectomy and demonstrated that the pain intensity and opioid requirement in patients that were administered IV ibuprofen were lower than in patients that were administered IV paracetamol. ^[4]^. Similarly, another study evaluated patients undergoing arthroscopic surgery and found that the VAS scores and rescue analgesia use in the group administered 800 mg IV ibuprofen were significantly lower than in patients that received ketorolac. ^[11]^ In contrast, there are studies suggesting that the administration of preemptive 400 mg IV ibuprofen does not have a significant effect on postoperative pain and opioid consumption. ^[16]^ Additionally, some other studies claim that 800 mg preemptive IV ibuprofen in elective umbilical hernia surgery does not have a significant effect on postoperative pain intensity. ^[17]^ Our findings supported the studies that suggest that 800 mg IV ibuprofen reduces postoperative pain and opioid use.

In a previous study evaluating patients undergoing LCC, it was reported that the administration of 50 mg dexketoprofen reduced postoperative pain and opioid use. ^[2]^ In another study conducted on the same patient group, it was found that 10 and 50 mg dexketoprofen reduced opioid consumption and there was no difference between the two doses. ^[3]^ Similarly, in various studies conducted with dexketoprofen, it was revealed that preoperative administration of dexketoprofen reduced postoperative opioid use by 36-50%. ^[4]^ Additionally, some other studies that evaluated patients undergoing various surgical operations demonstrated that postoperative pain levels and opioid use in patients treated with preoperative dexketoprofen were found to be significantly lower than those in control subjects. ^[9, 18, 19]^ Beside the studies evaluating ibuprofen, there are also numerous studies comparing the effects of dexketoprofen and other NSAIDs on postoperative pain. One of these studies compared dexketoprofen with lornoxicam and showed that the pain levels and opioid consumption were lower in patients treated with dexketoprofen. ^[19]^ By contrast, some longitudinal studies reported that dexketoprofen did not have any effect on postoperative pain. Among these studies, a study conducted in urological patients showed that dexketoprofen had no effect on postoperative pain and opioid consumption. ^[18]^ In our study, the findings indicated that preoperative IV dexketoprofen administration reduced postoperative pain and opioid consumption and the VAS scores were significantly lower in the dexketoprofen group than in the control group. Additionally, in a similar way to ibuprofen, opioid consumption at postoperative 6 and 24 hours was significantly lower in the dexketoprofen group compared to the control group. The reason as to why both drugs significantly reduced opioid consumption only within the first six hours could be due to their short half-lives (dexketoprofen: 2.7 hours, ibuprofen: 2.2 hours). Moreover, the administration duration of drugs significantly changes the level of maximum drug concentration in plasma (Cmax). The C max level is almost twice higher after the administration of IV ibuprofen for 30 min compared to 5-7 min. In our study, both drugs were administered for 30 min. Literature suggests that although the administration of these two drugs for 30 min provides effective analgesia, the Cmax level will be lower; therefore, a booster dose after postoperative 6 hours may further reduce opioid consumption. ^[20]^ Nevertheless, more comprehensive studies are needed to elucidate this phenomenon. Our study is of particular importance since, to our knowledge, it is the first study to compare the effectiveness of two drugs, ibuprofen and dexketoprofen, which belong to the same group. Our findings indicated no significant difference between the two drugs in terms of postoperative pain and opioid consumption. Although the VAS scores and opioid consumption were lower in the ibuprofen group, this difference was statistically insignificant. These findings implicate that the administration of these two drugs preoperatively has similar effects on postoperative pain and opioid consumption. Therefore, selection of these drugs should be performed by taking into consideration the potential side effects of these drugs in patients as well as the duration of the effects of the drugs and the cost of administration.

Non-steroidal anti-inflammatory drugs (NSAIDs) are known to cause various side effects such as gastrointestinal, renal, cardiovascular effects and bleeding tendency. ^[20]^ In contrast, ibuprofen weakly inhibits COX-1 and COX-2 and thus its side effects are less common than those of other NSAIDs. ^[20]^ In a study evaluating the safety of IV ibuprofen in 1751 patients, it was shown that IV ibuprofen was well tolerated, had few side effects, did not pose hematological and renal risks, and could be used safely in surgical operations. ^[21]^ Another study found that opioid-induced symptoms such as nausea and vomiting were less common in patients using ibuprofen after LCC surgery. ^[1]^ In a similar way to ibuprofen, dexketoprofen has also been shown to have fewer side effects and fewer opioid-induced symptoms such as nausea and vomiting^[21, 22]^. In our study, both drugs were well tolerated by the patients and no drug-related side effects were observed in any patient. Although opioid-induced symptoms such as nausea, vomiting, and dry mouth were seen in more patients in the control group, there was no significant difference between the control group and the treatment groups. Based on the findings of our study, we suggest that the reduction of opioid consumption by these two drugs will reduce the frequency of potential opioid-induced side effects in the patients. Further comprehensive studies with larger patient series are needed to substantiate our findings.

### Limitations

Our study was limited due to the fact that Bispectral Index Monitoring (BIS) could not be performed and thus the anesthetic depth of the patients could not be monitored, mainly due to the high cost of BIS administration. Additionally, hemodynamic changes occurring during the intraoperative period were controlled using fentanyl and thus patients were more exposed to the risk of side effects of opioids.

## CONCLUSION

The findings showed that preoperative IV ibuprofen and dexketoprofen administration reduces pain and opioid consumption, particularly within the first 6 hours after LCC. It was also revealed that both drugs are well tolerated and safe and can reduce the incidence of opioid-induced side effects and that there is no difference between the two drugs with regard to pain relief and opioid consumption.

## Data Availability

All relevant data are within the manuscript

## Availability of data and materials

The datasets used and analyzed in the current study are available from the corresponding author on reasonable request.

## REFERENCES

1. Ahiskalioglu EO, Ahiskalioglu A, Aydin P, Yayik AM, Temiz A. Effects of single-dose preemptive intravenous ibuprofen on postoperative opioid consumption and acute pain after laparoscopic cholecystectomy. Medicine (Baltimore). 2017;96:e6200.

2. Anıl A, Kaya FN, Yavaşcaoğlu B, Mercanoğlu E, Türker G, Demirci A. Comparison of postoperative analgesic efficacy of intraoperative single-dose intravenous administration of dexketoprofen trometamol and diclofenac sodium in laparoscopic cholecystectomy. J Clin Anesth. 2016;32:127–133.

3. Piirainen A, Kokki H, Immonen S, Matti E, Merja R H, Heidi H et al. A Dose-Finding Study of Dexketoprofen in Patients Undergoing Laparoscopic Cholecystectomy: A Randomized Clinical Trial on Effects on the Analgesic Concentration of Oxycodone. Drugs R D. 2015;15:319–328.

4. Gago Martínez A, Escontrela Rodriguez B, Planas Roca A, Martínez Ruiz A. Intravenous Ibuprofen for Treatment of Post-Operative Pain: A Multicenter, Double Blind, Placebo-Controlled, Randomized Clinical Trial. PLoS One. 2016;11:e0154004.

5. Carr DB, Goudas LC. Acute pain. Lancet. 1999;353:2051–2058.

6. Buvanendran A, Kroin JS. Multimodal analgesia for controlling acute postoperative pain. Curr Opin Anaesthesiol. 2009;22:588–593.

7. Sinatra RS. Multimodal Management Of Acute Pain: The Role of IV NSAIDs2011.

8. Mitchell RW, Smith G. The control of acute postoperative pain. Br J Anaesth. 1989;63:147–158.

9. Hanna MH, Elliott KM, Stuart-Taylor ME, Roberts DR, Buggy D, Arthurs GJ. Comparative study of analgesic efficacy and morphine-sparing effect of intramuscular dexketoprofen trometamol with ketoprofen or placebo after major orthopaedic surgery. Br J Clin Pharmacol. 2003;55:126–133.

10. Le V, Kurnutala L, SchianodiCola J, Khaja A, Joel Y, Jean DE, et al. Premedication with Intravenous Ibuprofen Improves Recovery Characteristics and Stress Response in Adults Undergoing Laparoscopic Cholecystectomy: A Randomized Controlled Trial. Pain Med. 2016;17:1163–1173.

11. Uribe AA, Arbona FL, Flanigan DC, Kaeding CC, Palettas M, Bergese SD. Comparing the Efficacy of IV Ibuprofen and Ketorolac in the Management of Postoperative Pain Following Arthroscopic Knee Surgery. A Randomized Double-Blind Active Comparator Pilot Study. Front Surg. 2018;5:59.

12. Gozeler MS, Sakat MS, Kilic K, Ozmen O, Can A, Ince I. Does a single-dose preemptive intravenous ibuprofen have an effect on postoperative pain relief after septorhinoplasty? Am J Otolaryngol. 2018;39:726–730.

13. Ciftci B, Ekinci M, Celik EC, Ahmet K, Muhammet AK, Yavuz D, et al. Comparison of Intravenous Ibuprofen and Paracetamol for Postoperative Pain Management after Laparoscopic Sleeve Gastrectomy. A Randomized Controlled Study. Obes Surg. 2019;29:765–770.

14. Ekinci M, Ciftci B, Celik EC, Köse EA, Karakaya MA, Ozdenkaya Y. A Randomized, Placebo-Controlled, Double-Blind Study that Evaluates Efficacy of Intravenous Ibuprofen and Acetaminophen for Postoperative Pain Treatment Following Laparoscopic Cholecystectomy Surgery. J Gastrointest Surg. 2020;24:780–785.

15. Çelik EC, Kara D, Koc E, Yayik AM. The comparison of single-dose preemptive intravenous ibuprofen and paracetamol on postoperative pain scores and opioid consumption after open septorhinoplasty: a randomized controlled study. Eur Arch Otorhinolaryngol. 2018;275:2259–2263.

16. Southworth S, Peters J, Rock A, Pavliv L. A multicenter, randomized, double-blind, placebo-controlled trial of intravenous ibuprofen 400 and 800 mg every 6 hours in the management of postoperative pain. Clin Ther. 2009;31:1922–1935.

17. Sparber LS, Lau CS, Vialet TS, Chamberlain RS. Preoperative intravenous ibuprofen does not influence postoperative narcotic use in patients undergoing elective hernia repair: a randomized, double-blind, placebo controlled prospective trial. J Pain Res. 2017;10:1555–1560.

18. Esparza-Villalpando V, Pozos-Guillén A, Masuoka-Ito D, Gaitán-Fonseca C, Chavarría-Bolaños D. Analgesic efficacy of preoperative dexketoprofen trometamol: A systematic review and meta-analysis. Drug Dev Res. 2018;79:47–57.

19. Sivrikoz N, Koltka K, Güresti E, Büget M, Sentürk M, Özyalçın S. Perioperative dexketoprofen or lornoxicam administration for pain management after major orthopedic surgery: a randomized, controlled study. Agri. 2014;26:23–28.

20. Wolfe LS. Eicosanoids: prostaglandins, thromboxanes, leukotrienes, and other derivatives of carbon-20 unsaturated fatty acids. J Neurochem. 1982;38:1–14.

21. Moore RA, Barden J. Systematic review of dexketoprofen in acute and chronic pain. BMC Clin Pharmacol. 2008;8:11.

22. Tuncer S, Pirbudak L, Balat O, Capar M. Adding ketoprofen to intravenous patient-controlled analgesia with tramadol after major gynecological cancer surgery: a double-blinded, randomized, placebo-controlled clinical trial. Eur J Gynaecol Oncol. 2003;24:181–184.

